# cg05883128 (DDX60) and NR3C2 (CD4 T-cells and B-cells) to Be the Genetic Roots of Systemic Lupus Erythematosus

**DOI:** 10.1101/2024.01.16.24301351

**Authors:** Zhengjun Zhang

## Abstract

**Motivation:** Systemic lupus erythematosus (SLE) is an autoimmune disease and a long-term condition affecting many body parts. Autoimmune diseases are affecting more people for reasons unknown, and the causes of these diseases remain a mystery.

**Method:** A newly introduced robust competing (risk) max-logistic regression classifier that can simultaneously perform subtype clustering and classification for disease diagnoses and predictions becomes a new hope to solve the mystery. We use this method in the study to discover critical DNA methylation CpG sites and genome genes, which lead to the highest accuracy and interpretability.

**Results:** The DNA methylation CpG site cg05883128 (DDX60) and gene NR3C2 are essentially responsible for SLE development. They can lead to 100% prediction accuracy together with a miniature set of other CpG sites and genes, respectively. cg05883128 (DDX60) reveals the LSE mechanism affecting many body parts. NR3C2 in CD4 T cells and B cells behaves reversely, leading to the cause of LSE and explaining the mechanism of the autoimmune disease.

**Conclusions:** This work represents a pioneering effort and intellectual discovery in applying the max-logistic competing risk factor model to identify critical genes for LSE, and the interpretability and reproducibility of the results across diverse populations suggest that the CpGs and DEGs identified can provide a comprehensive description of the transcriptomic features of SLE. The practical implications of this research include the potential for personalized risk assessment, precision diagnosis, and tailored treatment plans for patients.

## 1 Introduction

Systemic lupus erythematosus (SLE) is a long-term condition affecting many body parts —joints, skin, kidneys, blood cells, brain, heart, and lungs. It occurs when a person’s body’s immune system attacks her tissues and organs (autoimmune disease). The global SLE newly diagnosed population is estimated to be around 400,000 annually, and most of these people are women [1]. In the one year in review 2023 [2], the authors reviewed 93 published papers in 2022 and concluded new results and data have improved the understanding of SLE, although further studies and research are needed to improve the knowledge we have on this complex disease. Upon reviewing 120 published articles, the authors in [3] concluded that more robust immunological biomarkers are needed to better understand disease progression in individuals with SLE, including non-organ-specific SLE biomarkers and organ-specific SLE biomarkers. Since no single biomarker can be sensitive and specific enough for SLE, multiple biomarkers combined through mathematical models may be a good idea for assessing SLE. Moreover, advanced computational methods are required to analyze large datasets and discover novel biomarkers. This paper is not going to further conduct a literature review, and the readers are referred to two excellent review papers [2,3]. This work aims to fill in the gap discussed in [3].

The significant contributions of this paper are four-fold. 1) a miniature set of genes is identified at the genomic level to lead to 100% accuracy. This set of genes is mathematically and biologically interpretable via different cohort studies and ethnicities. 2) a miniature set of CpGs is identified at the DNA methylation level to lead to 100% accuracy. This set of CpGs shows universality and why SLE is so complex at methylation levels. 3) The DNA methylation CpG site cg05883128 (DDX60) and gene NR3C2 are essential and responsible for SLE development. 4) This paper finds NR3C2 in CD4 T-cells and B-cells behaves reversely compared to NR3C2 in whole blood and CD4+, which causes a Simpson’s paradox and the genetic discovery of the long puzzled autoimmune disease (LSE). 5) The Simpson’s paradox further leads to a new autoimmune disease theory and sheds light on many other new biological studies and discoveries.

## 2 Method

We apply the newly proven max-linear competing logistic regression classifier method to the confirmed LSE and healthy controls classifications. The new method is very different from other classical statistical and modern machine learning methods, e.g., random forest, deep learning, and support vector machine [4]. In addition, the new method has enhanced the interpretability of results, consistency, and robustness, as shown in the literature in studies of COVID-19 and biomarkers of several types of cancers [4-7]. This section briefly introduces the necessary notations and formulas for self-containing due to the different data structures used in this work. This new innovative approach can be classified as either an AI or machine learning algorithm. However, this new approach has an explicit formula and is interpretable.

Suppose *Y*_i_ is the *i*th individual patient’s LSE status (*Y*_i_ = 0 for LSE-free, *Y*_i_ = 1 for confirmed) and 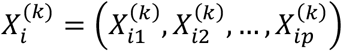, *k* = 1, …, *K* are the CpG beta values or gene expression values, with *p* = # *of* CpG sites or genes in this study. Here, *k* stands for the *k* th type of beta (expression) values drawn based on *K* different biological sampling methodologies. Note that most published works set *K* = 1, and hence the superscript (*k*) can be dropped from the predictors. In this research paper, *K* = 1, for DNA methylation data, *K* = 3, for gene expression data, and as we have three datasets analyzed in Section 3. Using a logit link (or any monotone link functions), we can model the risk probability 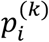 of the *i*th person’s infection status as:

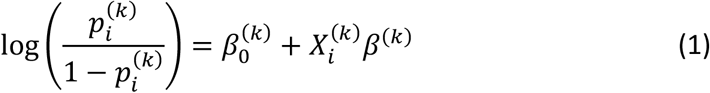

or alternatively, we write

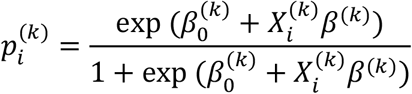

where 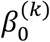 is an intercept, 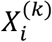 is a 1 x *p* observed vector, and *β*^(k)^ is a *p* x 1 coefficient vector which characterizes the contribution of each predictor (a CpG site or a gene, in this study) to the risk.

Considering the complexity of LSE, it is natural to assume that the epigenetic structures have subtypes and subtypes can be different. Suppose that all subtypes of LSE may be related to *G* groups of CpG sites (genes):

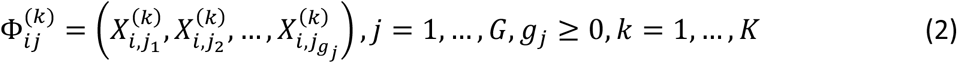

where *i* is the *i*th individual in the sample, and *g*_*j*_ is the number of CpG sites (genes) in *j*th group. The competing (risk) factor classifier is defined as:

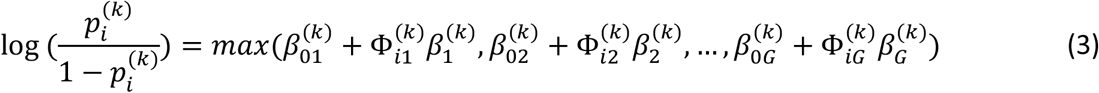

where 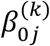s are intercepts, 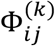 is a 1 x *g*_*j*_ observed vector, and 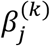 is a *g*_*j*_ x 1 coefficient vector which characterizes the contribution of each predictor in the *j* group to the risk.

**Remark 1**. *In (3)*, 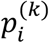 *is mainly related to the largest component* 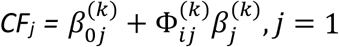, …, *G, i*.*e*., *all components compete to take the most significant effect*.

**Remark 2**. *Taking* 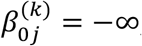, *j* = 2, …, *G, (3) is reduced to the classical logistic regression, i*.*e*., *the classical logistic regression is a special case of the new classifier. Compared with black-box machine learning methods (e*.*g*., *random forest, deep learning (convolutional) neural networks (DNN, CNN)) and regression tree methods, each competing risk factor in (3) forms a clear, explicit, and interpretable signature with the selected CpG sites (genes). The number of factors corresponds to the number of signatures, i*.*e*., *G* . *This model can be a bridge between linear models and more advanced machine learning methods (black box) models. However, (3) retains interpretability, computability, predictability, and stability properties. Note that this remark is similar to Remark 1 in Zhang (2021) [6]*.

We have to choose a threshold probability value to decide a patient’s class label in practice. Following the general trend in the literature, we set the threshold to be 0.5. As such, if 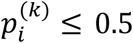, the *i*th individual is classified as being disease-free; otherwise, the individual is classified as having the disease.

Zhang (2021) [6] introduced a new machine learning classifier, the smallest subset and smallest number of signatures (S4), for *K* = 1. We extended the S4 classifier from *K* = 1 to *K* = 3 as follows:

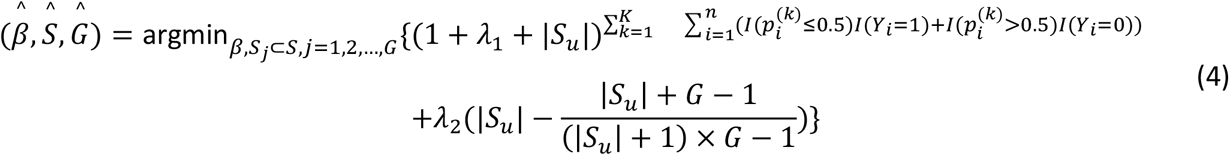

where *I*(. ) is an indicative function, 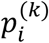 is defined in Equation (3), *S* = {1,2,…, *n*} is the index set of all CpG sites (genes), 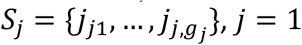, …, *G* are index sets corresponding to (2), *S*_u_ is the union of {*S*_*j*_, *j* = 1,…, *G*}, |*S*_u_ | is the number of elements in *S*_u_, *λ*_1_ ≥ 0 and *λ*_2_ ≥ 0 are penalty parameters, and 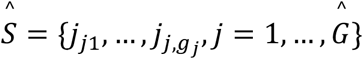 and *Ĝ* are the final CpG set (gene) selected in the final classifiers and the number of final signatures.

**Remark 3**. When the S4 classifier leads to 100% accuracy, the bioequivalence and DNA methylation (genome) geometry space can be established, which is a unique property established in (4) that does not appear in other classifiers in the literature [5].

**Remark 4**. *The case of K* = 1 *corresponds to the classifier introduced in Zhang (2021) [6]. The case of K* = 1 *and λ*_2_ = 0 *corresponds to the classifier introduced in Zhang (2021) [4]*.

**Remark 5**. *The computational details are referred to [7]*.

## 3 Data Descriptions, Results, and Interpretations

### 3.1 The data

We apply the optimization equation (4) to eight gene expression datasets (DEGs): GSE81622 **[8]** had Platform GPL10558: Illumina HumanHT-12 V4.0 expression beadchip. In this dataset, PBMC of 30 SLE patients, including 15 with LN (SLE LN+) and 15 without LN (SLE LN-), and 25 normal controls (NC) using HumanHT-12 Beadchips and Illumina Human Methy450 chips, were performed whole genome transcription. GSE97263 **[9]** had Platform GPL16791: Illumina HiSeq 2500 (Homo sapiens), mRNA extraction from isolated blood CD4+ T cells from 14 SLE with active disease, 16 SLE with less active disease and 14 controls. GSE177029 (two datasets FPKM and count) **[10,11]**, with Patforms (1) GPL24676:Illumina NovaSeq 6000 (Homo sapiens), examed 8 placentas from systemic lupus erythematosus and 8 from normal full-term pregnancies. GSE50722 **[12]** had Platform: GPL570 [HG-U133_Plus_2] Affymetrix Human Genome U133 Plus 2.0 Array with 61 SLE patients and 20 healthy controls. GSE144390 **[13]** had GPL6244 [HuGene-1_0-st] Affymetrix Human Gene 1.0 ST Array [transcript (gene) version] with three SLEs and three healthy controls. We included one DNA methylation dataset (CpGs): GSE82218 [8] with Platform PL13534 Illumina HumanMethylation450 BeadChip (HumanMethylation450_15017482). GSE4588 **[14]** (two datasets CD4 T cells and B cells sorted from total PBMC) has Platform: GPL570 [HG-U133_Plus_2] Affymetrix Human Genome U133 Plus 2.0 Array. CD4 T and B cells were sorted by flow cytometry from PBMC of patients with SLE, RA and healthy controls. GeneChip® Human genome U133 Plus 2.0 arrays were hybridized in monoplicates and the genes differentially expressed among the three groups of patients were identified using ANOVA tests with corrections for multiple comparisons. There are seven SLEs, nine healthy controls with B cells sorted, and nine SLEs and ten healthy controls with T cells sorted.

### 3.2 DEGs analysis

Following the computational procedure described in the earlier work [4-7], from 47322, 58037, 127550, 19943, 33298, and 54675 genes or gene IDs, six genes were identified to lead to 100% accuracy in six datasets GSE81622, GSE97263, GSE 177029, GSE50722, GSE144390, GSE4588. Table 1 lists the classifiers and the coefficients of the sites.

**Table 1.**
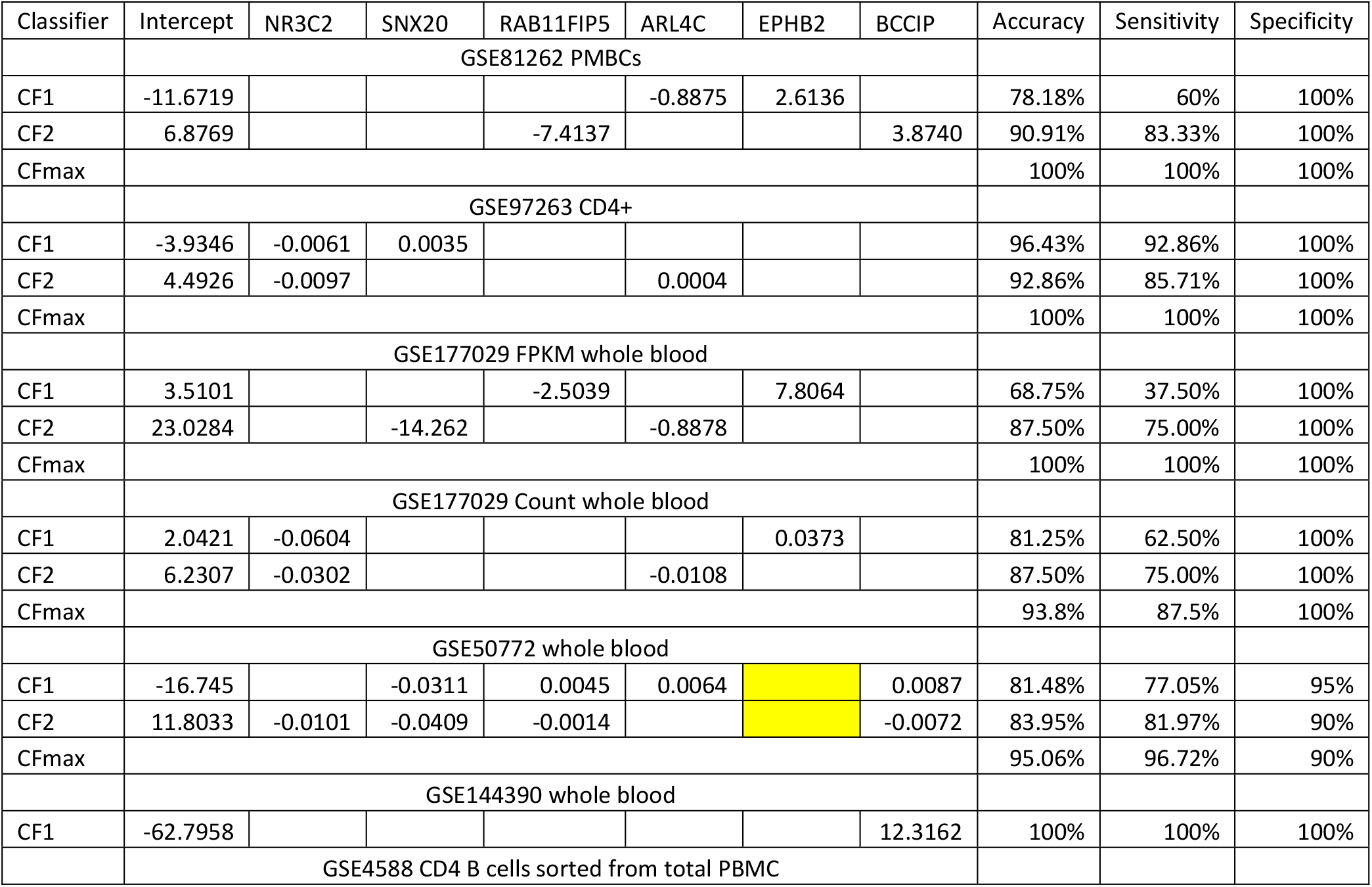
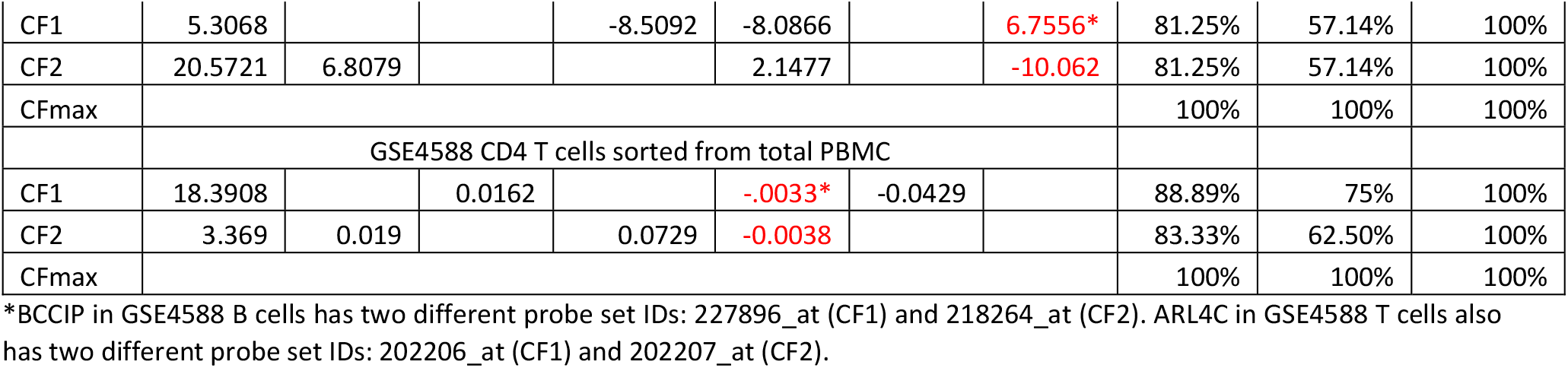
Performance of individual classifiers and combined max-competing classifiers using blood sampled data GSE81622, GSE97263, GSE177029, GSE50772, GSE144390, GSE4588 to classify SLE patients and non-SLE patients into their respective groups. CF1, 2 are two different classifiers. The numbers are fitted coefficient values.

In the table, each CFi is expressed as (using GSE81262 CF2 as an example):

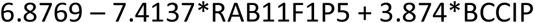

The six genes in genecords.org have the following descriptions. NR3C2 (Nuclear Receptor Subfamily 3 Group C Member 2) is a Protein Coding gene. Diseases associated with NR3C2 include Pseudohypoaldosteronism, Type I, Autosomal Dominant and Hypertension, Early-Onset, Autosomal Dominant, With Severe Exacerbation In Pregnancy. Among its related pathways are Gene expression (Transcription) and ACE Inhibitor Pathway, Pharmacodynamics. SNX20 (Sorting Nexin 20) is a Protein Coding gene. Diseases associated with SNX20 include Immunodeficiency 73A With Defective Neutrophil Chemotaxis And Leukocytosis and Inflammatory Bowel Disease.RAB11FIP5 (RAB11 Family Interacting Protein 5) is a Protein Coding gene. Diseases associated with RAB11FIP5 include Neonatal Lupus Erythematosus and Mucocutaneous Leishmaniasis. Among its related pathways are wtCFTR and delta508-CFTR traffic / Generic schema (norm and CF). Gene Ontology (GO) annotations related to this gene include small GTPase binding and gamma-tubulin binding. An important paralog of this gene is RAB11FIP1. ARL4C (ADP Ribosylation Factor Like GTPase 4C) is a Protein Coding gene. Diseases associated with ARL4C include Vulvar Leiomyosarcoma. Among its related pathways are NR1H2 and NR1H3-mediated signaling and ESR-mediated signaling. Gene Ontology (GO) annotations related to this gene include GTP binding and alpha-tubulin binding. An important paralog of this gene is ARL4A. EPHB2 (EPH Receptor B2) is a Protein Coding gene. Diseases associated with EPHB2 include Bleeding Disorder, Platelet-Type, 22 and Prostate Cancer/Brain Cancer Susceptibility. Among its related pathways are GPCR Pathway and EPH-Ephrin signaling. Gene Ontology (GO) annotations related to this gene include transferase activity, transferring phosphorus-containing groups and protein tyrosine kinase activity. An important paralog of this gene is EPHB1. BCCIP (BRCA2 And CDKN1A Interacting Protein) is a Protein Coding gene. Gene Ontology (GO) annotations related to this gene include RNA binding and kinase regulator activity.

Notably, except GSE50772 and GSE4588, all coefficients associated with NR3C2 and RAB11FIP5 are negative, which means increasing their expression values will decrease the risk of SLE symptoms. On the other hand, all coefficients associated with EPHB2 and BCCIP are positive, which means decreasing their expression values will decrease the risk of SLE symptoms. Two other genes do not have uniform coefficient signs, which complicates the SLE symptoms, leading to SLE subtypes, i.e., CF1 and CF2 can subdivide the patients into three subtypes in each dataset. Note that the three datasets were collected from UK (GSE97263), and China (two others). The subtypes can be different from different geographical regions. We can have a Venn diagram, as shown in Figure 1.

**Figure 1.**
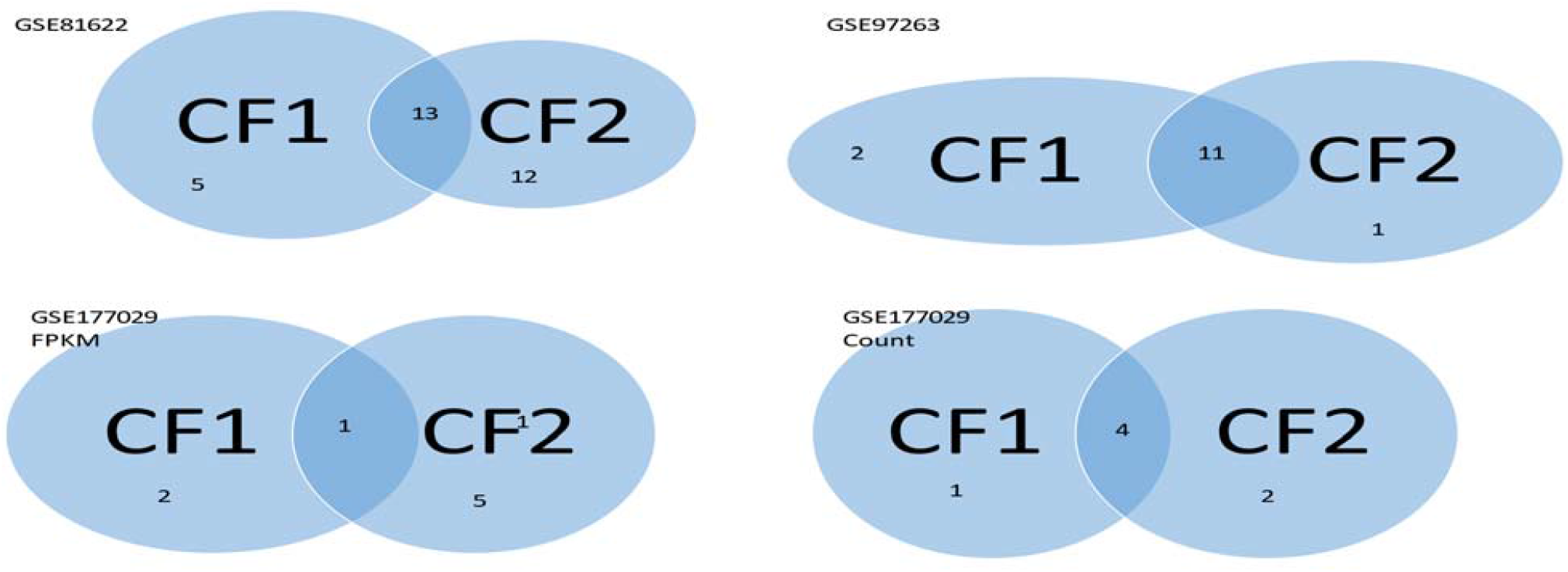
Venn diagrams are established from individual classifiers for each dataset.

Using GSE81622 as an example. The SLE patients are divided into three subtypes. Type 1 contains 5 patients who can only be detected by CF1; Type 2 contains 12 patients who can only be detected by CF2; Type 3 contains 13 patients whom both CF1 and CF2 can detect. Other Venn diagrams are interpreted similarly.

We note that GSE177029 contains two types of data: FPKM values and raw counts. It contains 127550 ensembl transcript (ENST) IDs. For illustration, we have GSE81622 containing 47322 ILMN IDs, and GSE97263 contains 58037 ensembl gene (ENSG) IDs. Some ENSG IDs can have multiple ENST IDs, which makes multiple solutions possible. Nevertheless, we still reached 100% accuracy with FPKM data.

Figure 2 presents risk probabilities evaluated for each individual in four datasets.

**Figure 2.**
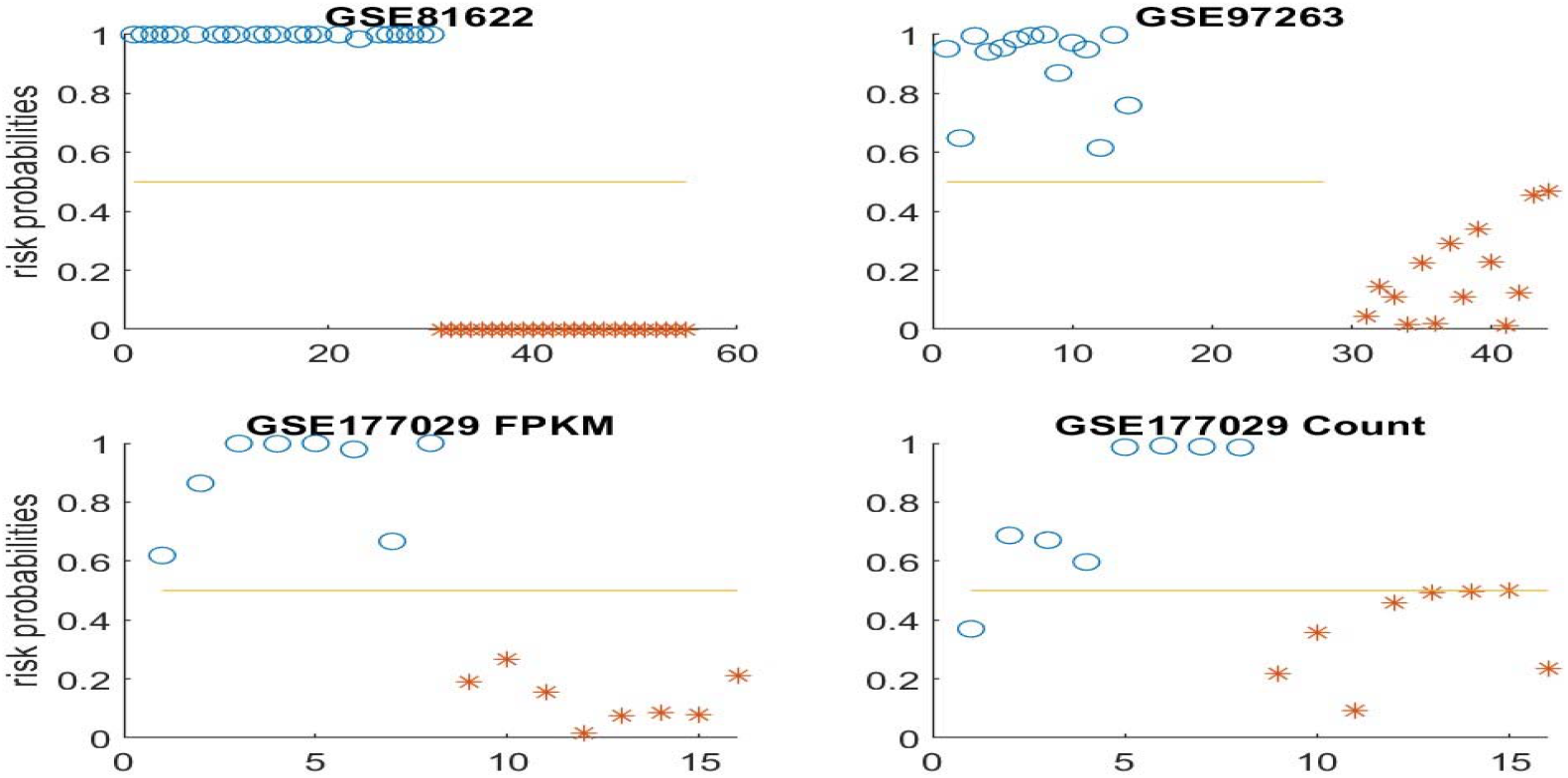
Risk probabilities for all individuals (SLE and controls). Blue circles correspond to SLE patients, while asters are for controls.

Figure 3 presents gene-gene interaction clinical view plots for GSE81622.

**Figure 3.**
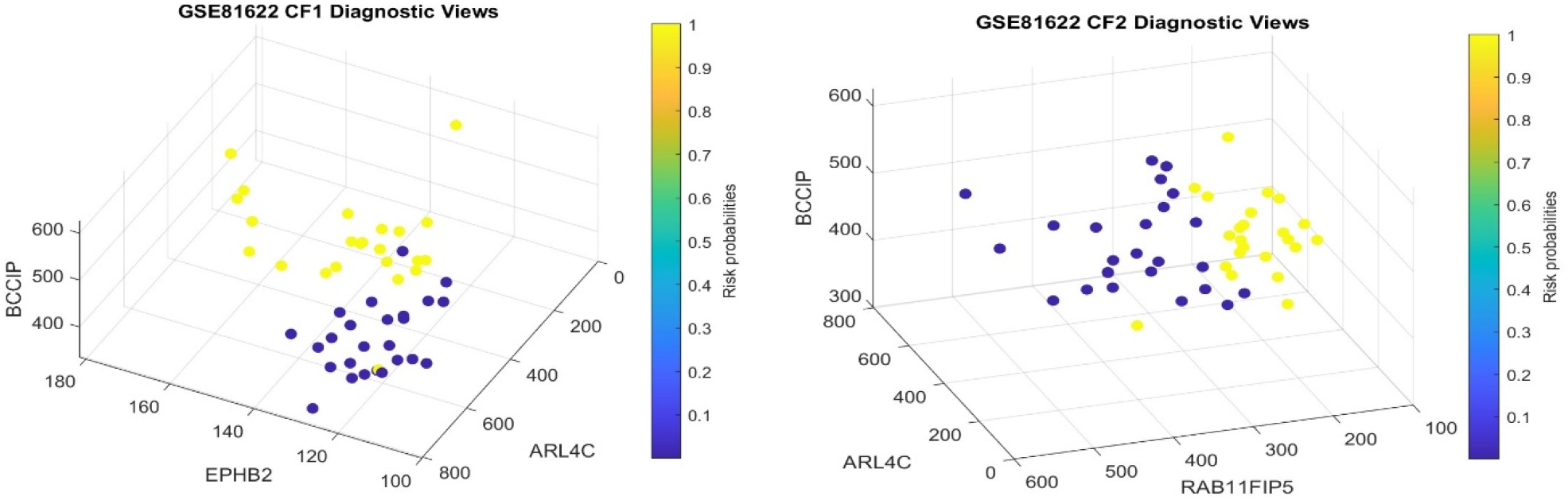
Clinical view of GSE81622.

Figures 2 and 3 demonstrate that the identified genes show the greatest separability between SLEs and controls, and these genes can be treated as critical biomarkers.

In GSE50772, there are only 19943 genes recorded. The gene EPHB2 was not found in the data. Nevertheless, the remaining five genes still led to an accuracy of 95.06%, a sensitivity of 96.72%, and a specificity of 90%. However, the coefficients associated with the genes RAB11FIP5 and BBCIP in GSE50772 are inconsistent with the signs in other datasets, making the interpretations of gene expression levels to the disease status not straightforward. Due to the missing EPHB2, its functions have to be allocated to other genes, and as a result, the signs of other genes may be affected, which does not disclose the truth. Such a phenomenon reveals that gene-gene interactions play pivotal roles in disease development, and finding driver genes is crucial to success.

In GSE4588, there are 54675 probe set IDs. Among them, (1,3,1,3,4,3) probe set IDs correspond to genes (NR3C2, SNX20, RAB11FIP5, ARL4C, EPHB2, BCCIP), respectively. In the footnote of Table 1, we listed the probe set IDs linked to ARL4C and BCCIP. The probe set IDs for NR3C2, SNX20, RAB11FIP5, EPHB2 used in Table 1 are 205259_at, 228869_at, 210879_s_at, 230088_at, respectively. At first glance, we see that the coefficient signs associated with genes in GSE4588 are reversed from the signs in other datasets, and we may conclude Simpson’s paradox exists in cell-level gene expression analysis. Note that in GSE4588, the expression values between CD4 T cells and B cells within several probe set IDs are significantly different. Figure 4 illustrates the differences. We can immediately see that DEGs at cell levels can be significantly different. As a result, it is not appropriate to put them in one model. Nevertheless, the genes identified for other cohorts still work perfectly with this cohort, indicating that these genes represent genetic insights into SLE. Likewise, the interpretations of gene functions in developing LSE have to consider its interactions with other genes and subtypes jointly.

**Figure 4.**
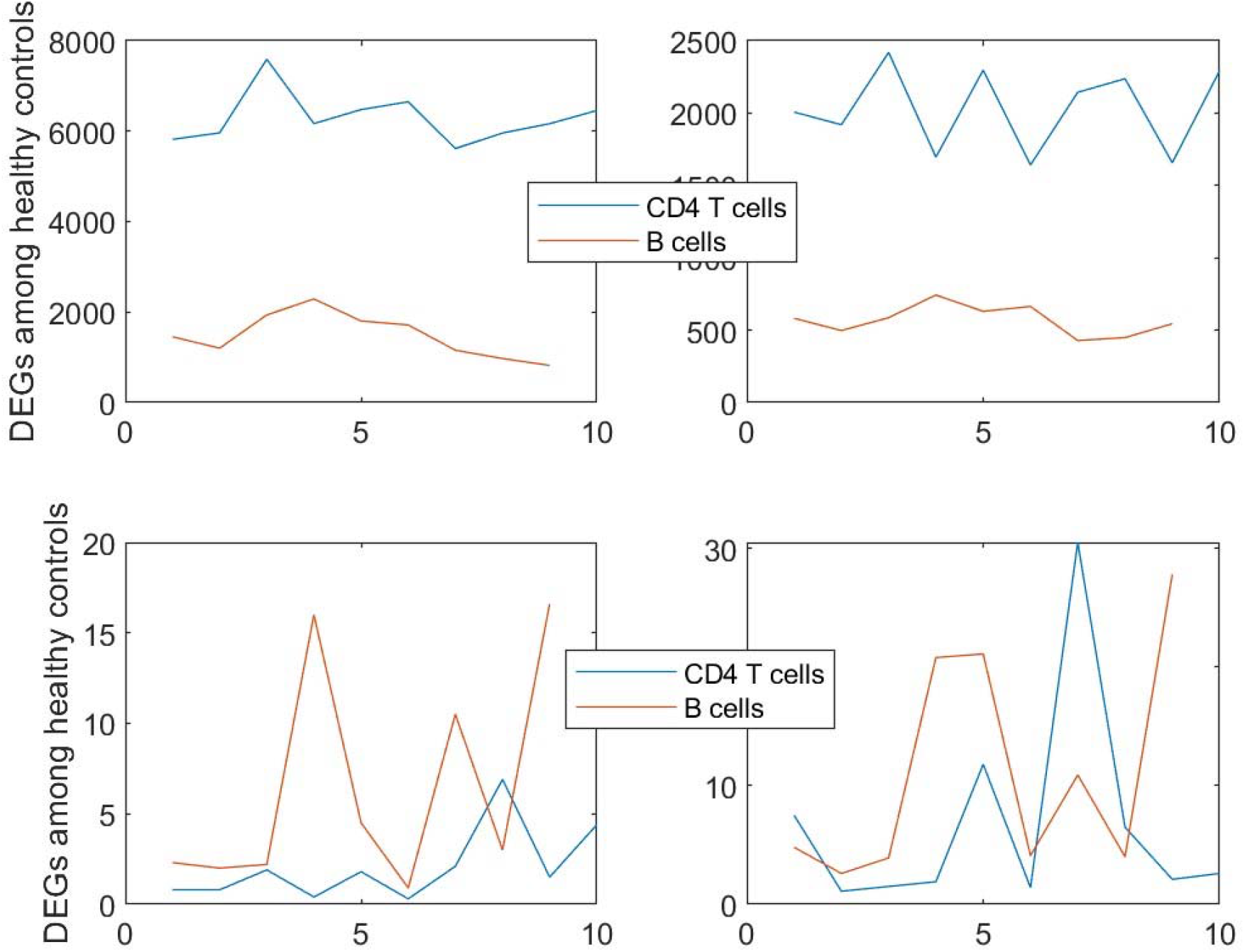
Illustrations of DEGs of four probe set IDs (genes) within healthy controls.

SLE has been regarded as an autoimmune disease, but its cause hasn’t been discovered in the literature. From our results in Table 1, we can hypothesize the following theory.

***Theory***: The cause of SLE is the increase of NR3C2 expressions in CD4 T-cells and B-cells.

### 3.3 DNA methylation analysis

From 485,577 methylation sites, cg05883128 (gene DDX60) was identified to lead to 100% accuracy in classifying SLE patients and non-SLE patients into their respective groups together with another single gene. Table 2 lists the classifiers and the coefficients of the sites.

**Table 2.**
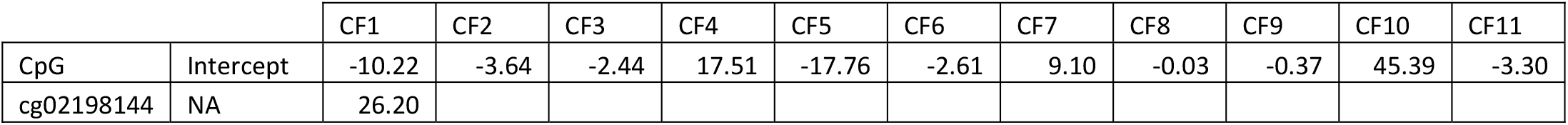
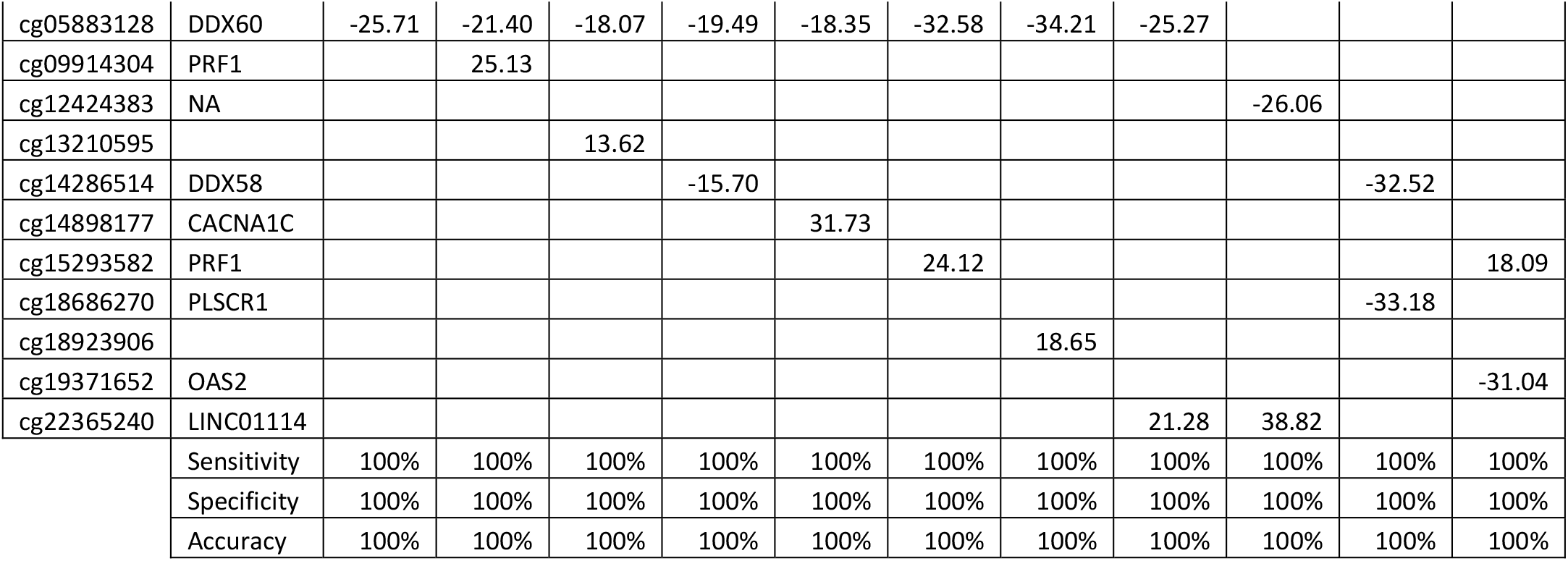
Performance of individual classifiers and combined max-competing classifiers using blood sampled DNA methylation data GSE82218 to classify SLE patients and non-SLE patients into their respective groups. CFi stands for an individual classifier. The numbers are fitted coefficient values.

From genecards.org, we can find that DDX60 (DExD/H-Box Helicase 60) is a protein coding gene. Diseases associated with DDX60 include Plantar Wart and Lip Cancer. Gene Ontology (GO) annotations related to this gene include nucleic acid binding and hydrolase activity. An important paralog of this gene is DDX60L. DDX58 RIGI (RNA Sensor RIG-I) is a Protein Coding gene. Diseases associated with RIGI include Singleton-Merten Syndrome 2 and Singleton-Merten Dysplasia. Among its related pathways are DDX58/IFIH1-mediated induction of interferon-alpha/beta and SARS-CoV-2 Infection. An important paralog of this gene is IFIH1. PLSCR1 (Phospholipid Scramblase 1) is a Protein Coding gene. Diseases associated with PLSCR1 include Hepatitis B and Influenza. OAS2 (2’-5’-Oligoadenylate Synthetase 2) is a Protein Coding gene. Diseases associated with OAS2 include Microphthalmia With Limb Anomalies and Tick-Borne Encephalitis. Among its related pathways are Interferon gamma signaling and Antiviral mechanism by IFN-stimulated genes. PRF1(Perforin 1) is a Protein Coding gene. Diseases associated with PRF1 include Hemophagocytic Lymphohistiocytosis, Familial, 2 and Aplastic Anemia. Among its related pathways are Granzyme Pathway and TCR signaling in naïve CD4+ T cells. Gene Ontology (GO) annotations related to this gene include calcium ion binding and wide pore channel activity. An important paralog of this gene is C6. CACNA1 (Calcium Voltage-Gated Channel Subunit Alpha1 C) is a Protein Coding gene. Diseases associated with CACNA1C include Timothy Syndrome and Long Qt Syndrome 8. Among its related pathways are DREAM Repression and Dynorphin Expression and TCR Signaling (Qiagen). Gene Ontology (GO) annotations related to this gene include enzyme binding and monoatomic ion channel activity. An important paralog of this gene is CACNA1D.

LINC01114 (Long Intergenic Non-Protein Coding RNA 1114) is an RNA Gene affiliated with the lncRNA class. Diseases associated with LINC01114 include Childhood Ependymoma.

Table 2 shows universality and complexity. On the one hand, the CpG site cg05883128 (gene DDX60) shows its coefficient signs are all negative, which indicates the cause of SLE could be cg05883128 CpG site much under-methylated, and this site can be a target curable medical treatment DNA methylation site. On the other hand, cg05883128 can combine and interact with eight other CpG sites to reveal SLE disease, which greatly increases the SLE complexity. In the combination classifiers CF1-3, CF5-8, increasing the methylation level of cg05883128 and decreasing the methylation level of the other CpGs can benefit pretecting SLE. It is interesting that cg14286514 (DDX58) can benefit the SLE status via increasing ite methylation level.

In addition, CF9, CF10, and CF11 are not related to cg05883128 (gene DDX60). Given their 100% accuracy, it is clear SLE has complex subtypes at DNA methylation levels.

Notably, the gene PRF1 has two CpGs involving the development of SLE. LINC01114 can interact with cg05883128 (gene DDX60) and cg12424383 for the development of SLE. Besides cg05883128 (gene DDX60), cg14286514 (DDX58) can be a key site for SLE development if it is under-methylated.

More importantly, Table 2 reveals why SLE can affect many parts of a person’s body as there are many gene (site) combinations, i.e., each can affect one part.

## 5 Discussions and Conclusions

### 5.1 Discussions

SLE is a complex disease, though not life-threatening. Given it can affect many parts of the body, its indirect cause of death can be an issue. Understanding the LSE disease and its development can help treat patients with multiple complex symptoms.

Among all genes identified in this paper, RAB11FIP5, DDX58, EPHB2 appeared in the list of hundreds of genes in [15]. We note that our critical gene detection method is significantly different from the literature approach. Our method focuses on site-site interaction, site-disease subtype interaction, gene-gene interaction, and gene-disease subtype interaction. As a result, the CpGs and genes identified in this paper can be interpreted using their fitted coefficients (signs) and their combinations with other sites and genes. Equation (4) outperformed AI, machine learning algorithm, and probabilistic algorithm in a COVID-19 study in [16] (Section 4.3).

In [2], the gene ITGA8 was discussed; see also [17]. However, this gene was not identified in our computation. It is worth studying further. In [9,18-19], epithelial stromal interaction 1 (EPSTI1) is associated with SLE. In [20-21], EPSTI1 and interferon (IFN)-alpha-inducible protein 27 (IFI27), were found related to SLE. IFI27 and EPSTI1 were found to have pivotal links to SARS-CoV-2 in [22]. Such a coincidence calls attention to COVID-19 infection-related SLE as so many people have been infected with COVID-19.

Besides genes reported earlier, this set of genes can also be insightful: MGC40489, IL18RAP, NUAK1, TOP1, MRPS18B, AL390026.1, ARPC4, LINC02762 as they also led to high and significant accuracy in our computations.

Many LSE research results at the genomic level have been published in the literature. These published results explored the pathological causes of SLE from various aspects. Due to study methodology limitations, some of the published results can hardly be cross-validated from cohort to cohort. Our work at the genomic level was a comprehensive study with nearly perfect performance. We didn’t find any other method that led to 100% accuracy in the literature, not even to mention interpretability. Many studies focused on only a single cohort whose representativeness cannot be assessed.

The dataset GSE4588 led to a Simpson’s paradox: using gene NR3C2 as an example. At the overall genomic level, the higher the DEGs, the better protection of LSE. On the other hand, at both CD4 T cells and B cell levels, the lower the DEGs, the better the protection of LSE. This phenomenon calls for further profound studies of how different cell types interact, e.g., cell clustering and classifications and identifying significant clusters. Also, the data process and quality have to be controlled.

We now discuss the most significant difference between our approach and the literature approach in finding critical genes. Much attention has been paid to the individual effects of every single gene in the literature due to the study design and available analysis methods. Our approach is jointly studying site-site, gene-gene, and gene-subtype interactions, which were largely missed in the literature. We can see from Tables 1-2 that the effects of each gene depend on other genes in the combinations. As a result, our findings of interaction effects can be the key to discovering the cause of SLE.

Many published results studied the functional effects of genes (including IFI27 and EPSTI1) based on single gene expression value changes. They lack interaction effects study, mainly due to the limitations of study methods. As a result, they lack accuracy and may not really be useful. Using the gene RAB11FIP5 as an example, in CF2 in Table 1 GSE81262, it must be jointly studied with another two genes EPHB2 and BCCIP, to fully understand its functional effects on SLE as its functional effects in CF1 with GSE177029 are significantly different.

Our results are nearly perfect, with some cohort studies with 100% accuracy and others with 95% or higher accuracy. In some scenarios, such nearly perfect results can be considered too good to be true. In our earlier work [16], we argued that the traditional cross-validation method is not applicable to our model (3). Instead, we apply cohort-to-cohort cross-validation in our earlier and present work. We used a driver gene dataset to demonstrate the superiority of our model (3) compared to those algorithms built for AI, machine learning, and deep learning. We found that our results are with better precisions, and more importantly, our results are interpretable [7, 22].

### 5.2 Conclusions

The pathological knowledge of the cause of the complex SLE is still unknown until now. The new findings of cg05883128 (DDX60) can shed light on new research and treatment plans. NR3C2 and RAB11FIP5 can be applied in medical treatment and drug development.

The theory of the cause of SLE is due to the increase of NR3C2 expressions in CD4 T-cells and B-cells will shed light on biological studies. The theory will completely change the ways genetic, cell, immunological studies on diseases are done.

## Acknowledgments

In memory of Kathy Chow Hoi-mei for playing the role of Zhou Zhiruo in “Heaven Sword and Dragon Sabre”. Chow had suffered from SLE disease and died on December 11, 2023, which motivated this research.

## Data Availability and Supplementary materials

The datasets are publicly available. The data links are stated in Section Data Description. Computing outputs are in a supplementary file available online https://pages.stat.wisc.edu/∼zjz/SLEmarkers.zip during the review process, and the file will be submitted to the publisher after the paper has been accepted. The results presented in this paper are all verifiable by simply checking the Excel sheets and formulas in the file.

## Author Contributions

Zhengjun Zhang is the sole author with 100% contributions to the article.

## Statement of ethics

The authors conducted research based on published work. The new research does not need IRB approval and a statement of ethics.

## Limitation statements

Compared with other studies the authors have conducted, the sample sizes in this study are relatively small. Given 100% accuracy in all datasets, which naturally and reasonably confirms the results are intrinsic. Of course, large-scale medical research can further verify the findings.

